# Outcomes Associated with Expanded Take-Home Eligibility for Outpatient Treatment with Medications for Opioid Use Disorder: A Mixed-Methods Analysis

**DOI:** 10.1101/2021.12.10.21267477

**Authors:** Oanh Kieu Nguyen, Scott Steiger, Hannah Snyder, Matthew Perrotta, Leslie W. Suen, Neena Joshi, Stacy Castellanos, Brad Shapiro, Anil N. Makam, Kelly R. Knight

## Abstract

**Background:** Access to medications for opioid use disorder (MOUD) in the U.S. is highly restricted. In March 2020, to reduce transmission of COVID-19, SAMHSA issued emergency regulations allowing up to two weeks of take-home doses for most patients.

**Objectives:** We evaluated the benefits and unintended consequences of these new regulations expanding take-home eligibility to inform MOUD policy post-pandemic

**Methods:** We conducted a mixed-methods evaluation of an opioid treatment program in San Francisco caring for a diverse, low-income urban population. We assessed clinic-level intake, retention, and take-home prescribing; individual-level acute care utilization and mortality; and patient/provider perceptions of benefits, harms and challenges of the new regulations.

**Results:** Clinic volume, intake and retention were largely unchanged after implementation of the new regulations, though the average monthly proportion of individuals receiving take-homes significantly increased from 31% to 47% (p<0.001). Among 506 established patients (≥90 days of care), the 10-month mortality was 2.7% among those who never received take-homes versus 3.2% among those newly started (p=0.79) and 0.8% among those with increases in take-homes (p=0.24). Individuals who never received take-homes had higher rates of emergency department visits (47.0%) and hospitalizations (19.7%) versus those with new starts (ED visits 29.2%, p<0.001; hospitalizations 14.3%, p=0.19) or increases in take-homes (ED visits 17.5%, p<0.001; hospitalizations 10.0%, p=0.02). Both patients and providers reported increased treatment flexibility, leading to increased engagement and stabilization.

**Conclusions:** Given the benefit and lack of appreciable harms, policymakers should consider extending expanded MOUD take-home eligibility after COVID-19, with careful monitoring for unintended outcomes.

## INTRODUCTION

Access to medications for opioid use disorder (MOUD) in the U.S. is highly restricted due to concerns about potential overdose, diversion, and misuse.^1^ Methadone and buprenorphine are effective, evidence-based life-saving medications for opioid use disorder (OUD) but buprenorphine can only be prescribed by specially licensed providers with a waiver from the Drug Enforcement Agency; methadone is further restricted to being administered or dispensed only at federally regulated opioid treatment programs (OTPs).^2, 3^ To receive take-home MOUD doses, the Substance Abuse and Mental Health Services Administration (SAMHSA) requires documentation that a patient is “responsible in handling narcotic medications” and that the rationale for take-homes considers the following stringent eight-point criteria: 1) absence of recent abuse of any drugs including alcohol; 2) regular clinic attendance; 3) absence of serious behavioral problems at clinic; 4) absence of known recent criminal activity; 5) stable home environment and social relationships; 6) adequate time-in-treatment for number of take-homes given (exempted for buprenorphine); 7) availability of safe storage for take-homes; and 8) benefit of decreased frequency of clinic attendance outweighs the potential risks of diversion.^4^

In March 2020, to mitigate the spread of SARS-CoV2-19 (COVID-19), SAMHSA issued emergency regulations that vastly expanded eligibility for take-home dosing, enabling OTPs to: 1) prescribe take-homes of up to 14 days for less stable patients and 2) prescribe up to 28-days of MOUD take-homes for all stable patients.^5^ There is currently limited evidence on the impact of these sweeping regulatory changes. The few published studies document marked increases in the number of take-home doses and proportion of patients receiving take-homes, low rates of self-reported diversion or misuse, and reductions in in-person dosing visits.^6-12^ Few robust data have been reported on initiation, retention, and stabilization in care due to expanded MOUD eligibility due to the new regulations, or on outcomes that might signal potential unintended harms, including mortality, acute care utilization, or reduction/revocation of take-homes (a proxy for diversion or misuse). One study of eight OTPs in Connecticut reported no increase in state-wide methadone-involved fatalities associated with an increase in take-home doses prescribed during a selected 40-day period after implementation of the new emergency regulations.^13^

We sought to understand the impact of the expanded eligibility for take-home MOUD dosing including benefits and unintended consequences, by assessing: 1) initiation and retention in OTP care pre- and post-new regulations; 2) mortality, acute care utilization, and revocation/reduction in take-homes among OTP patients receiving new take-homes in the 10 months after implementation of the new regulations; and 3) OTP patient and provider perspectives on benefits, harms, and potential challenges with implementation of the new regulations.

## METHODS

We conducted a mixed-methods evaluation at a hospital-affiliated OTP in San Francisco, California to triangulate the association of expanded eligibility on outcomes, benefits, and challenges of take-home MOUD dosing. The OTP cares for about 600 individuals per month; 95% are treated with methadone. The hospital is part of a safety-net health network of 12 primary care clinics and over 100 subspecialty clinics which serves a racially/ethnically diverse, low-income urban population and is linked by a common electronic health record (EHR) system.

For our mixed-methods approach, we used a convergent design to triangulate quantitative and qualitative findings.^14^ We designed the quantitative and qualitative evaluation arms in parallel to yield complementary findings. We then conducted the quantitative and qualitative evaluation arms concurrently, analyzed the data in each arm separately, and then converged the results for interpretation.

### Quantitative Evaluation

We conducted a retrospective analysis using EHR data from both the OTP (Methasoft) and health network (Epic) from January 2019-December 2020 for all individuals in OTP care.

#### Clinic-Level Outcomes

We assessed mean monthly clinic patient volume, mean monthly intakes and discharges, 60-day retention among new intakes, and the mean monthly proportion of clinic patients receiving any take-home doses pre-(January 2019-February 2020) vs. post-new regulations (March-December 2020).

#### Individual-Level Outcomes

We assessed outcomes among the cohort of individuals established in OTP care with a time-in-treatment greater than 90 days as of March 2020, and who therefore met time-in-treatment criteria for at least 1 take-home and had adequate time in treatment to meet the remaining eight-point criteria. We compared individuals who never received take-homes irrespective of regulatory changes to those who had any take-homes after the new regulations. We further categorized those with any take-homes by the first change in take-home status after new regulations: a) those newly started on take-homes; b) those with an increase in take-homes; and c) those with a decrease or no change in take-homes. We defined these groups to enable benchmarking outcomes among individuals with new starts of take-homes (our primary group of interest) to key reference groups likely to be at lower risk of adverse outcomes (individuals with any take-homes under the traditional regulations, subgroups b and c above) versus those at higher risk of adverse outcomes (those who never received take-homes regardless of the regulations). We opted to use this approach given the current lack of validated risk-adjustment approaches for adverse outcomes among individuals with OUD. We assessed outcomes over 10 months of follow-up through December 2020.

Key outcomes included mortality, emergency department (ED) visits and hospitalizations at the affiliated safety-net hospital, number of take-homes at the end of follow-up, reduction/revocations in take-home doses, avoided in-person visits, and unexcused clinic absences per month. We ascertained death from both EHRs and county department of public health records (updated from statewide vital records); cause of death was ascertained from medical examiner reports, death certificates, and/or EHRs when available.

#### Analytic Approach

We compared clinic-level outcomes before and after the new regulations by comparing mean monthly event rates using two-tailed t-tests. We compared individual-level outcomes using descriptive statistics, including the Kruskal-Wallis test to compare medians, and the z-test to compare proportions where applicable. An alpha level of p≤0.05 was considered statistically significant as per current academic convention.

### Qualitative Evaluation

#### Study Participants

We interviewed ten providers (physicians, nurse practitioners, nurses, and behavioral health counselors) and 20 patients from August-November 2020. We recruited providers through email, in-person outreach and snowball sampling to ensure diversity in roles. We recruited patients ≥18 years old in OTP care through provider referral and purposive sampling to obtain diverse perspectives. We aimed to recruit patients with diverse MOUD treatment experiences, duration, and treatment stability, and varied in COVID-19 exposure and risk.

#### Data Collection and Analysis

Semi-structured interviews focused on barriers to MOUD access and use, MOUD treatment experiences before and after COVID-related changes, and recommendations for care practices and regulatory changes post-COVID. We audio-recorded, transcribed, coded (Dedoose software) and content analyzed all interviews.^15^ The analysis used a thematic approach with simultaneous data collection and analysis based on modified grounded theory methodologies.^16^

The University of California San Francisco institutional review board deemed this study exempt from approval and informed consent as a program evaluation.

## RESULTS

The OTP had similar mean monthly total number of patients post-vs. pre-new regulations (571 vs 589, p<0.001), monthly numbers of new intakes (38 vs 42, p=0.46), discharges (28 vs 31, p=0.38), and 60-day retention rate among new intakes (63% vs. 69%, p=0.26). A mean of 31% of patients per month had any take-homes before vs. 47% per month (with an immediate, sharp increase) after the new regulations (p<0.001) (**Figure**).

**Figure.**
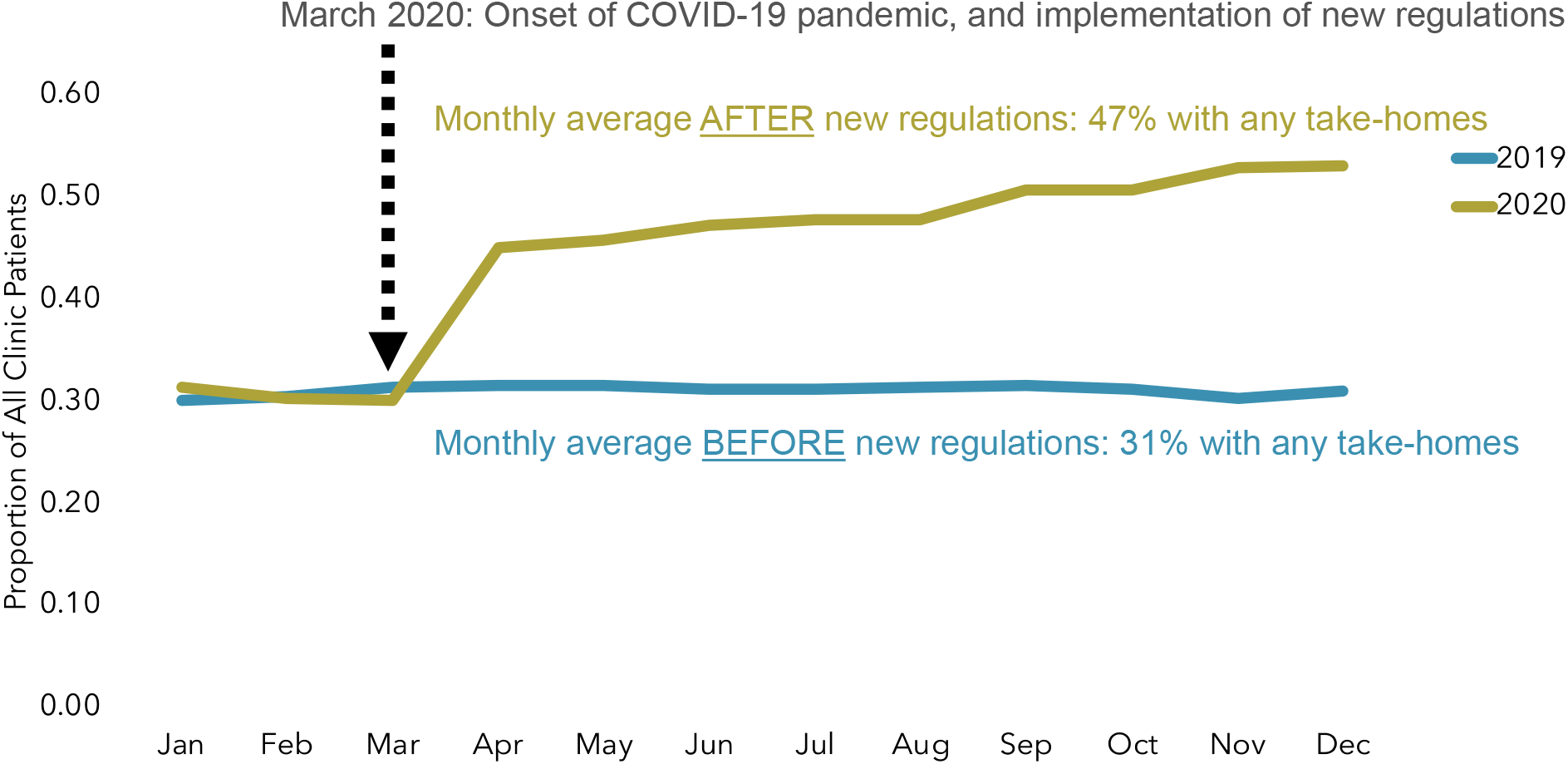
Average Monthly Proportion of OTP Patients with Any Take-Homes Before and After New MOUD Regulations. Abbreviations: MOUD, medications for opioid use disorder; OTP, opioid treatment program

### Outcomes Associated with Expanded MOUD Take-Homes

Among 506 individuals established in OTP care for >90 days, over one-third never had take-homes regardless of regulations (36.2%, n=183, **Table 1**). About one-third of established patients were newly started on take-homes (30.4%, n=154, **Table 1**) after the new regulations. One-third had take-homes before the new regulations (33.4%, n=169); among these individuals, after the new regulations most (71%, n=120) received an increase in the number of take-homes and 49 (29%, n=49) experienced a decrease or no change during the study period (in this subgroup, 63% were already receiving ≥2 weeks of take-homes, **Table 2**).

**Table 1.** Characteristics of Individuals Established in OTP Care* (n=506)

|  | Never Had Take-Homes (n=183) | Any Take-Homes After New Regulations† |  |  |
| --- | --- | --- | --- | --- |
|  |  | Newly Started (n=154) | Increase (n=120) | Decrease or No Change (n=49)‡ |
| Age, mean±SD | 47.5±11.2 | 52.8±11.7 | 41.7±14.0 | 54.3±12.1 |
| Male, % (n) | 76.5 (140) | 68.2 (105) | 59.2 (71) | 55.1 (27) |
| Race/ethnicity, % (n) |  |  |  |  |
| White | 51.4 (94) | 37.0 (57) | 40.8 (49) | 40.8 (20) |
| Black/African-American | 31.6 (57) | 33.1 (51) | 32.5 (39) | 26.5 (13) |
| Hispanic | 10.4 (19) | 20.1 (31) | 15.8 (19) | 18.4 (9) |
| Other | 7.1 (13) | 9.7 (15) | 10.8 (13) | 14.3 (7) |
| Health insurance, % (n) |  |  |  |  |
| Commercial | 0.6 (1) | 1.3 (2) | 3.3 (4) | 2.0 (1) |
| Medicare | 15.8 (29) | 23.4 (36) | 33.3 (40) | 24.5 (12) |
| Medicaid | 66.1 (121) | 55.8 (86) | 50.8 (61) | 53.1 (26) |
| Charity/self-pay/other | 17.5 (32) | 12.9 (20) | 12.5 (15) | 20.4 (10) |
| History of being unhoused§, % (n) | 53.0 (97) | 23.4 (36) | 3.3 (4) | 6.1 (3) |
| Time in treatment, years, median (IQR) | 2.0 (1.0-4.5) | 4.2 (1.4-9.7) | 5.1 (2.1-12.2) | 6.2 (3.3-11.2) |
| Time in treatment¶, % (n) |  |  |  |  |
| 91-180 days | 9.8 (18) | 7.1 (11) | 6.7 (8) | 2.0 (1) |
| 181-270 days | 10.4 (19) | 5.8 (9) | 2.5 (3) | 2.0 (1) |
| 271-365 days | 3.8 (7) | 4.6 (7) | 5.0 (6) | 4.1 (2) |
| 1-2 years | 25.1 (46) | 16.2 (256) | 10.8 (13) | 8.2 (4) |
| >2 years | 50.8 (93) | 66.2 (102) | 75.0 (90) | 83.7 (41) |

Abbreviations: IQR, interquartile range; OTP, opioid treatment program; SD, standard deviation
\* Individuals were considered 'established in OTP care' if they had a time in treatment of greater than 90 days as of March 2020. We selected this definition to limit analysis to individuals who met time-in-treatment criteria for at least 1 take-home and also had adequate time in treatment to potentially meet the remaining eight-point criteria under traditional regulations for take-homes, prior to the new regulations.
† Subgroup categories represent the first change in take-home dosing after implementation of the new SAMHSA emergency regulations, which allowed OTPs to: 1) issue take-homes of up to 14 days for less stable patients and 2) issue up to 28-days of take-homes for all stable patients. These new regulations replaced the prior eight-point criteria under the traditional regulations for take-homes.
‡ Included 19 individuals with an initial decrease in take-homes and 30 individuals with no-change in take-homes after the new regulations.
§ Defined as having housing status listed as 'homeless' as per either OTP or health system medical records.
¶ Categories correspond to time-in-treatment required per traditional regulations to receive a maximum of 2, 3, 6, 14, and 28 take-homes, respectively.

**Table 2.** Outcomes Associated with Changes in Take-Homes After New MOUD Regulations, Among Individuals Established in OTP Care*.

|  | Never Had Take-Homes (n=183) | Any Take-Homes After New Regulations† |  |  |
| --- | --- | --- | --- | --- |
|  |  | Newly Started (n=154) | Increase (n=120) | Decrease or No Change‡ (n=49) |
| Follow-up time, months, median (IQR) | 10 (7.7-10) | 10 (10-10) | 10 (10-10) | 10 (10-10) |
| Lost to follow-up, % (n) | 18.6 (34) | 4.5 (7) | 3.3 (4) | 6.1 (3) |
| Died, % (n) | 2.7 (5) | 3.2 (5) | 0.8 (1) | 4.1 (2) |
| Overdose § | 0.5 (1) | 0.6 (1) | 0.8 (1) | 4.1 (2) |
| Other | 0.5 (1) | 1.3 (2) | - | - |
| Unknown | 1.6 (3) | 1.3 (2) | - | - |
| Acute care utilization |  |  |  |  |
| Emergency department visits, % (n) | 47.0 (86) | 29.2 (45) | 17.5 (21) | 20.4 (10) |
| Median ED visits (IQR)¶ | 2 (1-3) | 1 (1-3) | 1 (1-2) | 3.5 (1-5) |
| Hospitalizations, % (n) | 19.7 (36) | 14.3 (22) | 10.0 (12) | 16.4 (8) |
| Median hospitalizations (IQR)¶ | 1 (1-1) | 1 (1-2) | 1 (1-1) | 1 (1-1) |
| Length of stay, days, median (IQR) | 4 (2-9.5) | 3.8 (2-5) | 2 (1.5-4) | 3 (2-6.5) |
| Any reduction in take-homes after initial change, % (n) | - | 46.1 (71) | 37.5 (45) | 16.4 (8) |
| Take-home doses, % (n) |  |  |  |  |
| Number of take-homes <u>before</u> new regulations |  |  |  |  |
| 0 | - | - | - | - |
| 1-5 (<1 week) | - | - | 37.5 (45) | 18.4 (9) |
| 6 (1 week) | - | - | 49.2 (59) | 18.4 (9) |
| 13 (2 weeks) | - | - | 13.3 (16) | 44.9 (22) |
| 27 (4 weeks) | - | - | 0.0 (0) | 18.4 (9) |
| Number of take-homes at the <u>end</u> of follow-up |  |  |  |  |
| 0 | - | 21.4 (33) | 7.5 (9) | 8.2 (4) |
| 1-5 (<1 week) | - | 65.6 (101) | 5.8 (7) | 14.3 (7) |
| 6 (1 week) | - | 9.7 (15) | 20.8 (25) | 14.3 (7) |
| 13 (2 weeks) | - | 3.6 (5) | 40.0 (48) | 42.9 (21) |
| 27 (4 weeks) | - | 0.0 (0) | 20.0 (24) | 20.4 (10) |
| Visits avoided/month, median (IQR) | 0 (0-0) | 2 (0-4) | 6 (4-13) | 6 (4-13) |
| Unexcused absences/month, median (IQR) |  |  |  |  |
| Before new regulations | 18% (6%-36%) | 11% (4-25%) | 0% (0-4%) | 0% (0-4%) |
| After new regulations | 25% (11-43%) | 11% (4-21%) | 4% (0-14%) | 8% (0-25%) |

Abbreviations: ED, emergency department; IQR, interquartile range; MOUD, medications for opioid use disorder; OTP, opioid treatment program
\* Outcomes were ascertained over a 10-month follow-up period after the new regulations, among all individuals established in OTP care (n=506). Individuals were considered 'established in OTP care' if they had a time in treatment of greater than 90 days as of March 2020. We selected this definition to limit analysis to individuals who met time-in-treatment criteria for at least 1 take-home and also had adequate time in treatment to potentially meet the remaining eight-point criteria under traditional regulations for take-homes, prior to the new regulations.
† Subgroup categories represent the first change in take-home dosing after implementation of the new SAMHSA emergency regulations, which allowed OTPs to: 1) issue take-homes of up to 14 days for less stable patients and 2) issue up to 28-days of take-homes for all stable patients. These new regulations replaced the prior eight-point criteria under the traditional regulations for take-homes.
‡ Included 19 individuals with an initial decrease in take-homes and 30 individuals with no-change in take-homes after the new regulations.
§ Defined as overdose from any or multiple illicit substances (including opioids) listed as any of the potential causes of death.
¶ Among those who had either an ED visit or hospitalization, respectively

Compared to those who never had take-homes, those with any take-homes after the new regulations were more likely to be female, non-white or to be housed. Those who never had take-homes had the lowest median time in treatment at 2.0 years (IQR, 1.0-4.5) with median time in treatment ranging from 4.2-6.2 years among those with any take-homes. (**Table 1**)

#### Mortality and Acute Care Utilization

Median follow-up time was 10 months overall (IQR 7.7-10), with the highest proportion lost to follow-up among those who never had take-homes (18.6%) versus other groups (range 3.3-6.1%, p=<0.005 for all comparisons). Mortality at 10 months was 2.7% (n=5) among those never receiving take-homes, versus 3.2% (n=5) for those newly started, 0.8% (n=1) for those with an increase in take-homes, and 4.1% (n=2) for those with no change or decrease (p>0.05 for all). Few deaths were attributed to drug overdose (2 total for the newly started and increased take-home groups combined).

Those who never had take-homes were far more likely to have ED visits (47.0%) and hospitalizations (19.7%) during follow-up, compared to those with any take-homes (ED visits, range 17.5-29.2%, p<0.001 for all comparison; hospitalizations, range 10.0-16.4, though only the comparison to those with an increase in take-homes was statistically significant, p=0.02). (**Table 2**).

#### Take-Homes at End of Follow-Up

At the end of follow-up, 21.4% of those newly started on take-homes had take-homes decreased to zero, versus 7.5% of those with an increase, and 8.2% of those with no initial change/decrease. (**Table 2**)

Additionally, at the end of follow-up, three-quarters (75.3%) of those newly started on take-homes had ≤1 week of take-homes, with only 3.6% receiving 2 weeks and 0% receiving 4 weeks of take-homes. Among those with increases in take-homes, 60.0% had ≥2 weeks of take-homes at the end of follow-up (versus 13.3% before the new regulations, p<0.001). (**Table 2**)

#### Provider Perceptions of Adverse Outcomes Associated with Expanded Take-Homes

Providers reported that adverse outcomes including overdose, diversion, and misuse of take-homes were minimal after new regulations despite initial concerns:

> *“What we haven’t seen are any like disaster stories…where someone got a few take-home doses of medication and like went and drank them all [and] overdosed. Like I think what it’s showing us is like, look, our clients, they’re legally and like very much emotionally, they’re all adults.”* [Provider]
>
> *“We have a bunch of patients who have been using stimulants for a long time who seem to have done really a fine job in terms of managing take-homes and like maybe they should have take-homes [after the pandemic].”* [Provider]

### Benefits of Expanded Take-Homes

#### Enhanced Stabilization of Opioid Use Disorder and Autonomy

Patients reported take-homes led to increased stabilization due to increased autonomy:

> *“[Take homes is] a lot of difference because I could take it [methadone] any time… when I need it…then I don’t got to get up to go out … be caught up in all that atmosphere…one of the dope areas…I don’t want to get caught up in, ‘Oh, I see you, hey, what’s going on…I got this [heroin] for you.’”* [Patient]
>
> *The whole thing [with getting take-homes] was just a game changer for me. Because there is no way I could have done it. I couldn’t go to school and carry a full load and be on methadone it would just not work.”* [Patient]

In parallel, expanded take-home dosing reduced mandatory in-person visits by a median of 6 visits per month among those with decreases/no change, 6 visits per month for those with increases in take-homes, and by 2 visits per month among those with new take-homes (**Table 2**). Additionally, unexcused absences increased for all groups after new regulations, with the notable exception of the newly started take-homes group, in whom unexcused absences remained stable (median 11%/month, IQR, 4-21%). (**Table 2**).

#### Enhanced Flexibility Around Treatment Decisions for Patients and Providers

Providers described using a case-by-case review approach that balanced risks and benefits and maintained flexibility in take-home decision-making:

> *[My counselor] seen…how hard I’ve been trying since I’ve actually been coming to the clinic, the transition that I’ve been making trying to elevate my life, so she didn’t want me to just turn around and say, “Well, I’ll go back on drugs, I won’t even have to come out here anymore.” [O]ne day I missed because I couldn’t get [to the clinic]…so she said, “I’m going to start to get you take-homes so you won’t have to come out here every day,”…So then she gave me another one, so for the last six months that’s the way it’s been for me*. [Patient]
>
> *“I think it’s just been a lot more honesty [about patient substance use] because clients know that we have the ability to give them take-homes for other reasons, like it’s just too burdensome for them to get to the clinic five days a week, perhaps they’re older and they have mobility issues…but they know that they don’t necessarily have to meet the same criteria, and so…I’ve had conversations with some clients that were very honest.”* [Provider]

Providers noted that despite increased flexibility, they were still cautious in prescribing take-homes, consistent with our finding 36.2% of patients were still dosing in-person daily even 10 months after implementation of new regulations:

> *“There’s two ways to go about it, right? You can either push it to the max and then dial back, or you implement and see the evidence and go forward, and we try to do the latter.”* [Provider]
>
> *“So, if I’m thinking, ‘Gosh, this guy’s like really sick [with COPD], and he’s on like five inhalers and still huffing and puffing.’ Then we’re going to present him to the team and be like, ‘I really want this guy to have take-homes. I feel like he could probably handle them. I feel like he has somewhere safe to put them. He is [currently] at a reasonably stable dose, so I don’t think we’ll have to be adjusting him all the time.’ So, we might start somebody out on just maybe weekend take-homes. […] He looks fine for a little while, then we’ll try like the next step up like if he like to show up on Monday, Wednesday and Friday, so we would have every other day off. And then you know kind of go from there. […] We have kind [of…] a blanket exception for COVID take-homes. But you know, it’s still risky so we’re weighing the risks.”* [Provider]

### Challenges with Implementation of Expanded Take-Home Dosing

Patients who did not receive take homes under new regulations perceived lack of transparency in the decision-making process, and drug exceptionalism, in which some drug use (e.g. stimulant use) was now tolerated for take-homes but not other types (e.g. benzodiazepines, opioids):

> *I understand nobody wants to lose their job but it really makes no sense that they make us [dose everyday] and expose us to people who are already stressed…it doesn’t make any sense – what’s the difference if I have something else in my system?* [Patient]
>
> *I’ll argue with them, too, like, “You show me in your book…you can’t just make up…stuff as you, as you go along because you think…I’m not going to tell you…that I’m doing it [fentanyl] or whatever…That’s, to me that’s not fair*. [Patienst]

## DISCUSSION

We found that new regulations expanding eligibility for take-home MOUD dosing were associated with a substantial and sustained increase in take-home prescribing without an associated increase in unintended harms (deaths and acute care utilization) among those newly started or those with new increases. Key benefits of expanded take-home MOUD dosing were perceived increased treatment flexibility with downstream improvement in care engagement and treatment stabilization. Despite expanded eligibility, providers remained judicious in their clinical decision-making, as evidenced by the high proportion of individuals still requiring daily in-person dosing, but was a potential source of frustration for patients not receiving take-homes given perceived lack of transparency.

Our findings are consistent with other recent studies which also found substantially increased take-home prescribing under the new regulations.^6-11^ One noteworthy finding was that our study site maintained similar rates of treatment initiation (in terms of unchanged new patient intakes) before and after the onset of the COVID-19 pandemic, which differs from national trends observed in Canada and the U.S. suggesting decreased availability of OTP treatment during COVID-19.^17^ In terms of individual clinical outcomes, few other studies to our knowledge have directly assessed outcomes among individuals newly started on take-homes under the new regulations, aside from one study from North Carolina which demonstrated no increase in self-reported diversion/misuse.^11^ Our finding that new take-homes were not associated with an increase in overall deaths mirrors the findings of a state-level study of methadone-involved fatalities in Connecticut, which found no increase in fatalities despite increased take-home prescribing across the state’s eight OTPs.^13^

Our findings have several implications for MOUD providers and policy makers. First, the lack of clearly and significantly increased harms of less restrictive take-home regulations in the context of considerable psychosocial stressors due to the COVID-19 pandemic – which we were unable to account for in our analyses, though separate data from our region showed a substantial increase in fatal drug overdoses during this time period – suggest an even larger potential magnitude of benefit of the new regulations than we present here.^18^ Second, the considerable increase in prescribing ≥2 weeks of take-home MOUD after the new regulations despite an extended time-in-treatment suggests potential underuse of take-homes under the previous regulations due to overly cautious prescribing. Keeping the current emergency regulations in place even after the COVID-19 pandemic may be one important means of mitigating barriers to MOUD access arising from underuse, and improving patient engagement in care.^19^ Finally, the majority of individuals newly started on take-homes received ≤1 week of doses and nearly half had dose reductions during the 10-months of follow-up, reflecting both the relapsing-remitting nature of chronic OUD and restraint and close monitoring by MOUD providers applying the new regulations, and argues against concerns of unrestrained MOUD prescribing by providers under the new regulations.

Our approach had some limitations. We did not have confirmatory information on methadone-related community overdoses nor information on MOUD diversion; however, we were able to ascertain deaths and acute care utilization from multiple sources, as well as dose reductions and revocations— strong proxies for diversion or misuse. Additionally, we were unable to risk-adjust analyses for individual risk factors given low event rates and the lack of a validated adjustment approach for adverse OUD outcomes. As a single-site study, we were unable to assess the impact of varying implementation across OTPs of new regulations on outcomes, an important topic for future study.^20^ Finally, we were unable to account for the concomitant effects of the ongoing COVID-19 pandemic and of the concurrent rising prevalence of unprescribed fentanyl use on our key outcomes. Both local and national data suggest increasing overdose deaths due to these factors during our study period.^18, 20-22^ As such, we anticipate even fewer adverse consequences of expanded eligibility for MOUD take-homes under the new regulations.

In conclusion, expanded eligibility for take-home MOUD dosing was not associated with an increase in unintended harms to patients, while improving engagement and flexibility in care. Given the considerable perceived benefit by both patients and providers, policymakers should consider extending regulatory changes beyond the end of the COVID-19 pandemic, with careful monitoring for unintended outcomes.

## Data Availability

All data produced in the present work are contained in the manuscript

## ACKNOWLEDGEMENTS

The authors thank Remy Hammel, Naomi López-Solano, and Miramar Kardouh for their contributions to data collection and Hannah Tierney for her contributions to study design. Study data were collected in part using REDCap electronic data capture tools hosted at UCSF. This work was supported by the California Health Care Foundation (G-31056 and G-31069). The sponsor had no role in the design and conduct of the study; collection, management, analysis and interpretation of the data; preparation, review or approval of the manuscript; no role in the decision to submit the manuscript for publication. The authors have no conflicts of interest to disclose.

## REFERENCES

1. National Academies of Sciences E, and Medicine; Health and Medicine Division; Board on Health Sciences Policy; Committee on Medication-Assisted Treatment for Opioid Use Disorder,. Medications for Opioid Use DIsorder Save Lives, Chapter 5: Barriers to Broader Use of Medications to Treat Opioid Use Disorder. National Academies Press; 2019 Mar 30.

2. Drug Addiction Treatment Act of 2000, https://www.congress.gov/bill/106th-congress/house-bill/2634 (2000). https://www.govregs.com/regulations/expand/title42_chapterI_part8_subpartC_section8.12

3. Opioid treatment program certification.

4. Substance Abuse and Mental Health Services Administration. Certification of Opioid Treatment Programs, 42 Code of Federal Reguations (CFR) 8.

5. FAQs: Provision of methadone and buprenorphine for the treatment of Opioid Use Disorder in the COVID-19 emergency. https://www.samhsa.gov/sites/default/files/faqs-for-oud-prescribing-and-dispensing.pdf

6. McIlveen JW, Hoffman K, Priest KC, Choi D, Korthuis PT, McCarty D. Reduction in Oregon’s Medication Dosing Visits After the SARS-CoV-2 Relaxation of Restrictions on Take-home Medication. Journal of Addiction Medicine. 9000;Publish Ahead of Printdoi:10.1097/adm.0000000000000812

7. Trujols J, Larrabeiti A, Sanchez O, Madrid M, De Andres S, Duran-Sindreu S. Increased flexibility in methadone take-home scheduling during the COVID-19 pandemic: Should this practice be incorporated into routine clinical care? J Subst Abuse Treat. Dec 2020;119:108154. doi:10.1016/j.jsat.2020.108154

8. Joseph G, Torres-Lockhart K, Stein MR, Mund PA, Nahvi S. Reimagining patient-centered care in opioid treatment programs: Lessons from the Bronx during COVID-19. J Subst Abuse Treat. Mar 2021;122:108219. doi:10.1016/j.jsat.2020.108219

9. Peavy KM, Darnton J, Grekin P, et al. Rapid Implementation of Service Delivery Changes to Mitigate COVID-19 and Maintain Access to Methadone Among Persons with and at High-Risk for HIV in an Opioid Treatment Program. AIDS Behav. Sep 2020;24(9):2469–2472. doi:10.1007/s10461-020-02887-1

10. Amram O, Amiri S, Thorn EL, Lutz R, Joudrey PJ. Changes in methadone take-home dosing before and after COVID-19. J Subst Abuse Treat. Jun 24 2021:108552. doi:10.1016/j.jsat.2021.108552

11. Figgatt MC, Salazar Z, Day E, Vincent L, Dasgupta N. Take-home dosing experiences among persons receiving methadone maintenance treatment during COVID-19. J Subst Abuse Treat. Apr 2021;123:108276. doi:10.1016/j.jsat.2021.108276

12. Munro A, Booth H, Gray NM, et al. Understanding the Impacts of Novel Coronavirus Outbreaks on People Who Use Drugs: A Systematic Review to Inform Practice and Drug Policy Responses to COVID-19. Int J Environ Res Public Health. Aug 11 2021;18(16)doi:10.3390/ijerph18168470

13. Brothers S, Viera A, Heimer R. Changes in methadone program practices and fatal methadone overdose rates in Connecticut during COVID-19. J Subst Abuse Treat. Apr 29 2021;131:108449. doi:10.1016/j.jsat.2021.108449

14. Creswell J. W., Plano Clark V.L. Chapter 3: Core Mixed Methods Designs. Designing and Conducting Mixed Methods Research. 3rd Edition ed. SAGE Publications; 2018.

15. Hsieh HF, Shannon SE. Three approaches to qualitative content analysis. Qual Health Res. Nov 2005;15(9):1277–88. doi:10.1177/1049732305276687

16. Corbin J., & Strauss A. Basics of qualitative research: Techniques and procedures for developing grounded theory. SAGE Publications, Inc; 2008.

17. Joudrey PJ, Adams ZM, Bach P, et al. Methadone Access for Opioid Use Disorder During the COVID-19 Pandemic Within the United States and Canada. JAMA Netw Open. Jul 1 2021;4(7):e2118223. doi:10.1001/jamanetworkopen.2021.18223

18. Appa A, Rodda LN, Cawley C, et al. Drug Overdose Deaths Before and After Shelter-in-Place Orders During the COVID-19 Pandemic in San Francisco. JAMA Netw Open. May 3 2021;4(5):e2110452. doi:10.1001/jamanetworkopen.2021.10452

19. Frank D, Mateu-Gelabert P, Perlman DC, Walters SM, Curran L, Guarino H. “It’s like ‘liquid handcuffs”: The effects of take-home dosing policies on Methadone Maintenance Treatment (MMT) patients’ lives. Harm Reduct J. Aug 14 2021;18(1):88. doi:10.1186/s12954-021-00535-y

20. Becker SJ, Garner BR, Hartzler BJ. Is necessity also the mother of implementation? COVID-19 and the implementation of evidence-based treatments for opioid use disorders. J Subst Abuse Treat. Mar 2021;122:108210. doi:10.1016/j.jsat.2020.108210

21. CDC Health Alert Network. Increase in Fatal Drug Overdoses Across the United States Driven by Synthetic Opioids Before and During the COVID-19 Pandemic December 2020;

22. American Medical Association. Issue brief: reports of increases in opioid-related overdose and other concerns during COVID pandemic. December 2020;

